# Analysis of Engagement with *CuidaTEXT*, a Text Message Intervention for Dementia Caregiver Support among Latinos

**DOI:** 10.1101/2023.12.06.23299630

**Authors:** Valerie Medina, Amber Watts, Monica Fracachán-Cabrera, Cameo Hazlewood, Mariana Ramirez-Mantilla, Eric D. Vidoni, Jaime Perales-Puchalt

## Abstract

**Objectives:** To assess the impact of a text message intervention, *CuidaTEXT*, on participant engagement and its subsequent associations with socio-demographic factors, acceptability measures, and clinical outcomes among Latino dementia caregivers.

**Methods:** *CuidaTEXT* is a six-month, bilingual, and bidirectional intervention. We enrolled 24 Latino caregivers in a one-arm feasibility trial. Participants received approximately one automatic daily text message and could engage with the intervention by texting specific keywords (e.g., STRESS to receive messages about stress-coping), and by chat-texting with a live coach. We used metrics and psychometric scales to quantify variables.

**Results:** Participants sent a total of 1,847 messages to *CuidaTEXT*. Higher engagement was associated with higher intervention satisfaction (r=0.6, p=0.007), as were several other acceptability outcomes. We found no associations between engagement with *CuidaTEXT* and sociodemographic or clinical outcomes (p>0.05).

**Conclusion:** Encouraging interaction with *CuidaTEXT* might lead to higher levels of satisfaction with the intervention. However, it might be that those who are highly satisfied, engage more with *CuidaTEXT*. Future research should determine the directionality of these associations.

**Clinical Implications:** Increasing opportunities to engage in text messaging may increase Latino caregivers’ satisfaction with caregiver support interventions.

## Introduction

Mobile health technologies (mHealth) have demonstrated significant potential in delivering support to family caregivers of individuals with Alzheimer’s disease and related dementias (ADRD). Family caregivers of individuals with ADRD often experience a disproportionate impact on their mental health, with higher rates of depression and other negative health outcomes.^1–8^ However, accessing caregiver support can be challenging for these individuals due to barriers, including time incompatibilities, transportation limitations, and limited intervention availability.^9^ To overcome such barriers, mHealth interventions, including telephone calls, apps, video-calls, and text messages, have surfaced as effective resources.

Latino family caregivers have higher rates of depression and less access to ADRD caregiver support interventions compared to non-Latino Whites.^10,11^ mHealth interventions may be accessible to many Latinos who are unable to attend in-person caregiver support. However, Latinos’ low use of most of these modalities would perpetuate their disparities in access to caregiving support.^12^ Text messaging is an ideal modality to deliver caregiver interventions to Latinos, as most Latinos own a cell phone and they use them for texting even more than non-Latino Whites.^13^ Text message interventions 1) are effective in treating various health disorders, 2) can be used anywhere at any time, 3) are more cost-effective compared to other delivery systems and 4) can be tailored to caregiver preferences and characteristics including language, culture and needs.^14–17^

Despite being one of the oldest mHealth modalities, the evidence of text messaging interventions for ADRD caregiver support is in its infancy. To our knowledge there is only one text message intervention to provide ADRD caregiver support, *CuidaTEXT.*^18,19^ This intervention specifically targeted Latino caregivers, and has shown promising feasibility, acceptability, and preliminary efficacy by providing crucial resources, information and emotional support to this specific population.^18^ While waiting to explore the fully-powered efficacy of this intervention, the potential of caregiver engagement with ADRD caregiver support text message interventions is still to be exploited.

This study aimed to explore participant engagement with a text message intervention for ADRD caregiver support, and its association with sociodemographic, acceptability and clinical variables. To achieve this aim, we analyzed data from our feasibility study.^18^ We hypothesized that a higher engagement with *CuidaTEXT* would be associated with increases in caregiver satisfaction with the intervention and decreases in depressive symptoms. A higher engagement with the program might help address a higher number of unmet needs and increase caregivers’ sense of support, which may, in turn, increase caregiver satisfaction with the intervention and decrease depressive symptoms.

## Methods

We present a secondary analysis of a one-arm pre-post-intervention trial design with assessments conducted at baseline and six months after intervention, in addition to text messaging metrics of engagement (e.g., number of messages, type of messages). We have described the details about the design, procedures, and participants of this study in a previous publication.^18^ In short, we enrolled 24 informal caregivers in August 2021 using a variety of recruitment techniques including research registries, community promotion, and advertisements in national organization registries and websites, outreach at health fairs, and word of mouth.

Eligibility criteria included speaking Spanish or English, being 18 or older, identifying as Latino, owning a cellphone with a flat fee, being able to read and write, and providing hands-on care for a relative with a clinical or research dementia diagnosis who also tested positive in a cognitive videocall/phone screener with the research team. For the current paper, we analyzed the data as a mixed-methods sequential study, in which qualitative data were used to define variables that were subsequently analyzed quantitatively. The quantitative method dominates, while the qualitative method is nested within. The quantitative method included descriptive statistics and correlations whereas the qualitative method included a qualitative descriptive approach. All study procedures were approved by the Institutional Review Board of the University of Kansas Medical Center (STUDY00144478). All participants gave written informed consent in their preferred language.

### Intervention

*CuidaTEXT* is a bilingual, six-month intervention tailored to caregiver needs via SMS text messages. *CuidaTEXT* is bidirectional, as participants receive scheduled text messages, but they can also text to receive on-demand messages. An in-depth description of the intervention and its development has been previously reported. ^18,19^ The intervention was designed from the beginning with and for Latino caregivers with the support of a team including bicultural, bilingual researchers, and informed by the Stress Process Framework and Social Cognitive Theory. ^20,21^ These messages include the identification of barriers to desired behaviors (e.g., problem solving, relaxation techniques, or exercising), setting of realistic goals, encouragement of gradual practice to increase healthy behaviors, integration of testimonials and videos to promote vicarious learning, integration of praise, social support, and education to increase dementia knowledge. *CuidaTEXT* includes approximately one scheduled daily automatic message (n=244 over six months) about logistics, dementia education, self-care, social support, end-of-life care, care of the person with dementia, behavioral symptoms, and problem-solving strategies.

Participants can also text two types of messages to receive on-demand assistance via: 1) up to 783 keyword-driven text messages providing on-demand help for the above topics; 2) live chat interaction with a coach from the research team for further help upon request. Before enrollment, staff mail participants a 19-page reference booklet summarizing the purpose and functions of the intervention in their preferred language.

### Assessment

The research team collected information from three sources: baseline survey, six-month follow-up survey, and metrics of text message engagement. The research team published the description of baseline characteristics of the sample previously.^18^ We also collected cell phone characteristics in the baseline survey, which include questions developed by the team about whether the participant was the account holder of the cellphone (yes/no), the self-reported number of SMS text messages sent and received by the participants per day previous to the program, whether participants share the cellphone with family members (yes/no), the frequency with which participants check their cell phones for messages (5-item Likert scale ranging from every 1-4 minutes to less than once a day), whether participants had ever signed up for SMS text message reminders (yes/no), and whether participants use Wi-Fi, data or none to access internet on their phones.

Variables included engagement with *CuidaTEXT*, sociodemographic characteristics, acceptability, and clinical characteristics:

<u>(1) Engagement with *CuidaTEXT*</u> included metrics on the number of each type of text messages sent by participants, which we categorized as total, keyword-driven (and their subgroups, prespecified during intervention development), and chat text (specified based on the qualitative analyses described in this manuscript). These data were obtained from a spreadsheet automatically downloadable from the SMS messaging software. These spreadsheets included information about the participant ID number, content of the message, date, and time the message was sent.
<u>(2) Sociodemographic characteristics</u> included the following binomial variables: gender (women/men), ever signing up for SMS text message reminders (yes/no), caregiver insurance status (yes/no), marital status (yes/no), and Spanish as preferred language (yes/no). We also included the following ordinal and continuous variables: age (in years), number of texts sent and received per day before the program, frequency of checking for text messages before the program, number of days a week participants saw their loved one with ADRD in person, financial inadequacy (5-point Likert scale ranging from very easy to very difficult), and the number of hours a day participants felt they were on caregiving duty.
(1) <u>(3) Acceptability outcomes</u> were based on previous text message health research ^22,23^ and were all collected in the follow-up survey. These outcomes included nine four-point Likert scale questions on satisfaction with *CuidaTEXT* and its components (Not at All-Extremely). Three additional four-point Likert scale questions asked about their perceived helpfulness of *CuidaTEXT* in: caring for the individual with dementia, caring for themselves, and enhancing their understanding of dementia (Not at All-A Lot).
(1) <u>(4) Clinical characteristics</u> included scales administered at baseline and follow-up, which we have described previously.^18^ These scales include the Neuropsychiatric Inventory– Questionnaire (caregiver distress; NPI-Q-D),^24,25^ Modified Caregiver Strain Index (CSI),^26^ Zarit Burden Interview-6 (ZBI-6),^27^ Positive aspects of caregiving scale (PAC),^28^ Epidemiology/Etiology Disease Scale (EEDS),^29,30^ Interpersonal Support Evaluation List (ISEL- 12),^31^ Coping Orientation to Problems Experienced Inventory (COPE-28),^32,33^ Center for Epidemiologic Studies Depression Scale (CES-D-10),^34,35^ Scale of Positive and Negative Experience (SPANE),^36,37^ Preparedness for Caregiving Scale (PCS),^38,39^ and Self-perceived health.^40^

### Analysis

We used descriptive statistics to summarize demographic, clinical and cellphone-related baseline characteristics of caregivers, and participants’ engagement. To analyze the chat text messages, we used a qualitative description methodology, which is a vehicle to present and treat research methods as living entities that resist simple classification, and thematic analysis methods, which emphasizes identifying, analyzing, and interpreting patterns of meaning within qualitative data.^41–43^ We coded the content of the spreadsheet by identifying codes. These codes were later categorized into domains. Two researchers (JPP and MFC) conducted the coding, contrasted, and resolved coding disagreements through discussion and consensus. We excluded in the qualitative analysis any chat text messages that related to reminders for the follow-up assessment.

We analyzed associations between participant engagement and the following variables: (1) sociodemographic characteristics, (2) baseline clinical characteristics, (3) acceptability outcomes, (4) change in clinical characteristics from baseline to follow-up. Since participant engagement was not distributed normally, we conducted non-parametric analyses in all cases. These included Wilcoxon Rank sum tests for binomial variables and Spearman correlations ordinal and quantitative variables. We used SPSS v20.0 for all calculations ^44^. The significance level was set at p < 0.05. Tables present the main findings whereas Appendices include findings from all analyses.

## Results

A total of 24 caregivers participated in this study. Among them, 20 (83%) were women, 10 (41.7%) were born in the USA, 6 (25.0%) were born in Mexico, and 8 (33.3%) were born in another Latin American country. An overview of participant characteristics is presented in Table 1. The average age of caregiver participants was 52.6 years (SD 13.2) and ranged from 26-81 years. Approximately half of the caregivers were married or had a partner 13 (54.2%). On average, caregivers had completed 14.7 (SD 3.8) years of education. Of the participants, 20 (83%) were adult children or child-in-laws of an individual with dementia, while 4 (16.7%) were spouses or partners. The average score for depressive symptoms using the CES-D-10 was 9.1 (SD 4.4). Most participants (75.0%) reported being the cellphone account holder, and they reported checking their phones at different intervals, with 41.7% checking every 5-59 minutes and 33.3% checking every few hours.

**Table 1.** Description of demographic, clinical and cellphone-related characteristics of the sample.

|  | Total (n=24) |
| --- | --- |
| Age in years, mean (SD) | 52.6 (13.2) |
| Women, % (n) | 83.3% (20) |
| Country of birth, % (n) |  |
| US, % (n) | 41.7% (10) |
| Mexico, % (n) | 25.0% (6) |
| Other, % (n) | 33.3% (8) |
| Years of education, m (SD) | 14.7 (3.8) |
| Spanish only as primary language, % (n) | 41.7% (10) |
| Relation to care recipient |  |
| Children, % (n) | 75.0% (18) |
| Children-in-law, % (n) | 8.3% (2) |
| Partner, % (n) | 16.7% (4) |
| CES-D-10 depression (range 0-30), mean (SD) | 9.1 (4.4) |
| Participant is account holder of cellphone, % (n) | 75.0% (18) |
| Number of text messages sent daily, median (IQR) | 10.0 (4.3-20.0) |
| Number of text messages received daily, median (IQR) | 17.5 (4.3-37.5) |
| Shares cellphone with family members, % (n) | 8.3% (2) |
| Frequency of checking for text messages |  |
| Every 1-4 minutes, % (n) | 12.5% (3) |
| Every 5-59 minutes, % (n) | 41.7% (10) |
| Every few hours, % (n) | 33.3% (8) |
| A couple of times a day, % (n) | 8.3% (2) |
| Less than once a day, % (n) | 4.2% (1) |
| Ever signed up for text message reminders, % (n) | 66.7% (16) |
| Uses Wi-Fi, data or non on their phone |  |
| Both, % (n) | 95.8% (23) |
| None, % (n) | 4.2% (1) |

Table 2 shows findings domains and examples from the qualitative data analysis of text messages sent by caregivers. This analysis yielded eight distinct domains. Domains included expressions of Gratitude, Acknowledgement, Logistics, Education, Behavior, Care, Caregiver, and Other.

**Table 2.** Domains abstracted from participants’ text chat engagements and representative excerpts.

| Domain | Excerpt |
| --- | --- |
| Gratitude | "Thank you for an incredible 6 months of support... I will miss you!", "Thank God and yourself for the help you provide!", "Thanks for the unconditional support. I am grateful for the daily texts to reinforce my caregiver role", "Hello. May I share the video link above on Alz/Dementia with my mother? It's so well-presented and quite informative", "Thank you very much. It's been very helpful to hear other talk about their experiences with their loved ones". |
| Acknowledgement | "When my mother is sad, I call her sister or friends in her home country. They talk to her about fun things they experienced together", "Thanks, I have called the Alzheimer's Association on 3 occasions these last 3 years", "I use my calendar to keep track of everything", "It's a blessing for me to take care of my mom. She sacrificed her whole life for her kids abs now I can do something for her - by taking care of her!", "When she does something wrong, it works to be patient and understand that it's not their fault", "My little dog, she is always with me. She makes my days less stressful". |
| Logistics | "I got the booklet. Have a nice day!", "BEHAVIOR and SUPPORT", "Alzheimer's Association in Spanish", "F", "[after texting several words] I am just testing out this program with what I know", "Can I recommend books that help me along my journey?", "Did the keyword not work because I didn't use upper case? I'll try again", "Am I texting to an individual or to a group?", "We're going to Mexico tomorrow, we have arranged to be able to receive |
|  | messages there, thanks!", "Yes, I am receiving the text messages (SMS). However, I cannot receive images (MMS), due to the settings of my plan", "I don't understand this message very well". |
| Education | "Is the treatment much varied among the different types?", "Is there a definitive exam/test to identify which type is affecting a patient?", "Could eating/hunger issues be dependent on the areas of the brain affected by the disease?", "What are the typical complications that people with Alzheimer's die of?", "Does anyone with Alzheimer's ever get better?", "I'd be interested in finding information about legal aspects". |
| Behavior | "Just brought Mom home from hospital and rehab after three weeks. She can no longer live alone so she's living with us. She said, 'I want to go home.' Over 30 times yesterday and started up again this morning. We try to change the subject to distract her, try to do an activity with her but she persists 'I want to go home!'", "I'm worried about my loved one's anger. It's hard to help him because he refuses help". |
| Care | "My mother won't stop scratching her head. I worried she'll break her skin", "I'd like to find a good daycare facility that has Spanish-speaking staff in our city", "What kind of stool or step can I get so my loved one can get into my car?", "My loved one's doctor is retiring so I'm looking for a doctor that is good with elderly patients with Alzheimer's Disease", "Please let me know when you're available to call me and help me fill the request for a respite grant for my husband", "My mother is the other way round. I need to know how to get her to stop taking so many showers". |
| Caregiver | "I'd like help explaining to my husband how tired I am from caring for my mother, and that we need to find help to be financially ready for when she gets worse. Are there social workers or specialists to guide couples about these |
|  | topics?", "I have not heard back from the respite grant to care for my husband", "Hi there, can you please remind me where I can find psychological help for myself?". |
| Other | "Merry Christmas to you!", "Happy Thanksgiving!", "Happy New Year to CuidaTEXT Support!", "[Responding to a scheduled question about relaxation needing to be practiced to learn the skill] Yes/No", "I do have a question regarding your study. May I ask the thesis of your research if it will not compromise your results?". |

Participants sent a total of 1,847 messages to *CuidaTEXT*. Engagement levels varied among participants, with two (8.3%) showing no engagement (no messages sent), three (12.5%) demonstrating low engagement (sent <10 messages), six (25.0%) displaying medium engagement (sent between10 and 49 messages), seven (29.2%) exhibiting high engagement (sent between 50 and 99 messages), and six (25.0%) demonstrating very high engagement (sent more than 100 messages). Table 3 shows the descriptive statistics of each type and domain of text messages.

**Table 3.** Description of participants’ engagement with the different types of messages.

|  | Median | Minimum | Percentile 25 | Percentile 75 | Maximum |
| --- | --- | --- | --- | --- | --- |
| Total number of messages | 51.0 | 0 | 14.3 | 97.5 | 392 |
| Total number of keywords | 14.5 | 0 | 5.0 | 30.8 | 309 |
| Total number of chat text messages | 30.5 | 0 | 5.0 | 52.0 | 177 |
| Keyword messages; Care domain | 3.0 | 0 | 2.0 | 7.0 | 80 |
| Keyword messages; Caregiver domain | 1.5 | 0 | 0.3 | 8.3 | 98 |
| Keyword messages; Support domain | 3.0 | 0 | 0.0 | 5.8 | 132 |
| Keyword messages; Behavior domain | 4.0 | 0 | 0.0 | 7.8 | 69 |
| Chat text messages; Gratitude domain | 9.0 | 0 | 1.5 | 17.3 | 144 |
| Chat text messages; Acknowledgement domain | 2.0 | 0 | 0.0 | 15.0 | 42 |
| Chat text messages; Logistics domain | 1.0 | 0 | 0.0 | 2.8 | 8 |
| Chat text messages; Education domain | 0.0 | 0 | 0.0 | 1.0 | 7 |
| Chat text messages; Behavior domain | 0.0 | 0 | 0.0 | 0.0 | 7 |
| Chat text messages; Care domain | 0.0 | 0 | 0.0 | 9.5 | 156 |
| Chat text messages; Caregiver domain | 0.0 | 0 | 0.0 | 0.0 | 3 |
| Chat text messages; Other domain | 1.0 | 0 | 0.0 | 1.8 | 2 |

Engagement with *CuidaTEXT* was generally not associated with the baseline sociodemographic or clinical characteristics (Table 4; Appendix 2). The few exceptions were associations with preferred language (Spanish-speakers were more engaged with total [mean rank difference=8.4; p=0.004] and keyword messages [mean rank difference=7.8; p=0.007]), and hours a day on caregiving duty (the more hours of caregiving, the more engaged with total [r=0.43; p=0.036] and chat text messages [r=0.58; p=0.003]).

**Table 4.** Association between engagement with CuidaTEXT and baseline sociodemographic and clinical characteristics.

|  | Total number of messages | Total number of keywords | Total number of chat text messages |
| --- | --- | --- | --- |
| Women (Mean rank difference) | -2.85 | -4.35 | 0.45 |
| Ever signed up for text message reminders (Mean rank difference) | -1.69 | 0.84 | -4.5 |
| Caregiver insurance (Mean rank difference) | -5.43 | -4.67 | -4.04 |
| Spanish as preferred language (Mean rank difference) | 8.40* | 7.81* | 5.54 |
| Age (Rho) | 0.19 | 0.07 | 0.24 |
| Years of education (Rho) | -0.25 | -0.08 | -0.36 |
| Number of text messages received daily (Rho) | -0.36 | -0.26 | -0.23 |
| Financial inadequacy (Rho) | 0.18 | 0.23 | 0.11 |
| Hours a day on caregiving duty (Rho) | 0.43* | 0.17 | 0.58* |
| NPI-Q Distress (Rho) | -0.076 | -0.070 | 0.021 |
| Caregiver Strain Index (Rho) | 0.224 | 0.085 | 0.365 |
| ZBI Burden (Rho) | -0.129 | -0.244 | 0.002 |
| CES-D-10 depression (Rho) | -0.14 | -0.18 | -0.07 |
| SPANE-Positive affect (Rho) | 0.32 | 0.33 | 0.19 |
| SPANE-Negative affect (Rho) | -0.31 | -0.09 | -0.23 |
| Perceived health (Rho) | 0.20 | 0.36 | -0.11 |

*Note.* Spearman's Rho: statistical dependence between the rankings of two variables; \* $p < 0.05$

Engagement with *CuidaTEXT* was significantly associated with several acceptability outcomes (Table 5; Appendix 3). For example, higher total message engagement was positively correlated with intervention satisfaction (r=0.6, p=0.007), participant perceptions that *CuidaTEXT* provided help for their loved one with ADRD (r=0.6, p=0.003), and increased understanding of ADRD (r=0.6, p=0.008). One of the few associations that did not reach statistical significance includes the correlation between total message engagement and participant perceptions that *CuidaTEXT* provided help for themselves as caregivers (r=0.3, p=0.173).

**Table 5.**
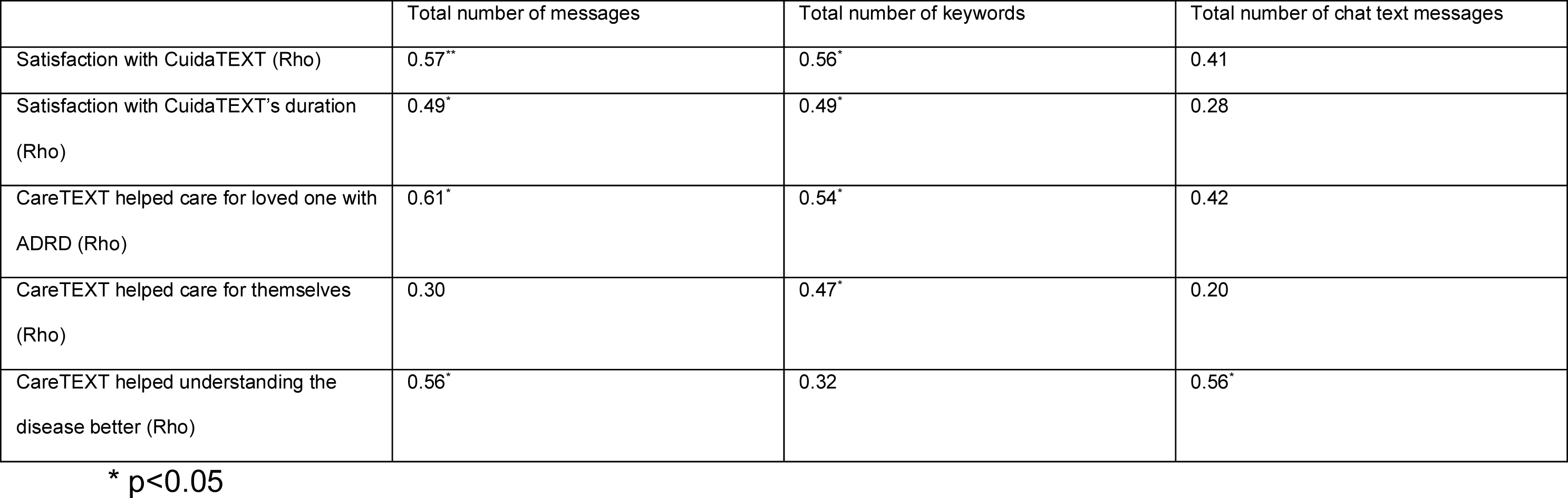
Association between engagement with CuidaTEXT and acceptability outcomes.

|  | Total number of messages | Total number of keywords | Total number of chat text messages |
| --- | --- | --- | --- |
| Satisfaction with CuidaTEXT (Rho) | 0.57** | 0.56* | 0.41 |
| Satisfaction with CuidaTEXT's duration (Rho) | 0.49* | 0.49* | 0.28 |
| CareTEXT helped care for loved one with ADRD (Rho) | 0.61* | 0.54* | 0.42 |
| CareTEXT helped care for themselves (Rho) | 0.30 | 0.47* | 0.20 |
| CareTEXT helped understanding the disease better (Rho) | 0.56* | 0.32 | 0.56* |

\* $p < 0.05$

Engagement with *CuidaTEXT* was generally not associated with the change in clinical outcomes from baseline to follow-up (Table 6; Appendix 4). For example, total message engagement with *CuidaTEXT* did not show a statistically significant correlation with changes in caregiver depressive symptoms (r=-0.0, p=0.922). Other examples that showed no statistically-significant correlation with total message engagement with *CuidaTEXT* include caregiver self-rated symptoms (r=-0.0, p=0.863) or burden (r=-0.1, p=0.765).

**Table 6.** Association between engagement with CuidaTEXT and baseline to follow-up change in clinical characteristics.

|  | Total number of messages | Total number of keywords | Total number of chat text messages |
| --- | --- | --- | --- |
| NPI-Q Distress (Rho) | 0.43 | 0.37 | 0.06 |
| Caregiver Strain Index (Rho) | 0.04 | -0.12 | 0.16 |
| ZBI Burden (Rho) | 0.07 | -0.03 | 0.13 |
| CES-D-10 depression (Rho) | -0.00 | -0.01 | -0.12 |
| SPANE-Positive affect (Rho) | -0.33 | -0.16 | -0.25 |
| SPANE-Negative affect (Rho) | 0.10 | -0.00 | 0.16 |
| Perceived health (Rho) | -0.04 | -0.03 | 0.04 |

\* $p < 0.05$

## Discussion

This study aimed to explore participant engagement with a text message intervention for ADRD caregiver support among Latinos, and its association with demographic characteristics, and acceptability and clinical outcomes. This represents the first text message intervention for caregiver support of individuals with ADRD among Latinos or any other ethnic group, and has shown high levels of acceptability, and preliminary efficacy.^18,19^ For this reason, exploring the associations between the level of engagement with the intervention and these outcomes is crucial. We hypothesized that higher engagement with CuidaTEXT would provide higher levels of support, which in turn would lead to higher levels of satisfaction and decreases in depressive symptoms. Our findings support the satisfaction but not the depressive symptoms hypothesis. In general, engagement with *CuidaTEXT* was associated with acceptability outcomes, but not demographic and clinical characteristics, nor change in clinical outcomes.

Participants in our study who engaged more with the program reported higher levels of satisfaction with the intervention, along with several other acceptability outcomes. Findings align with a study that found that texting frequency was associated with relational satisfaction among college students.^45^ Similarly, in non-texting interventions, greater engagement and adherence to interventions (e.g., cognitive behavioral therapy for eating disorders) have shown to increase therapeutic alliance or connection between patient and therapist.^46,47^ A potential interpretation of our findings is that engagement with *CuidaTEXT* may have resulted in greater emotional closeness with the intervention and, in turn, led to positive acceptability outcomes.

The frequency of engagement was generally not associated with change in clinical outcomes. This finding is not consistent with our hypothesis, where we expected more improvement in depressive symptoms among those with higher levels of engagement. For example, a study where 28 consecutive patients with obsessive compulsive disorder were treated with exposure and response prevention found that compliance with in-session and homework exposure instructions was significantly related to posttreatment symptom severity.^48^ Another study where patients with depression received cognitive behavioral therapy found that those who did more homework improved more than those who did little or no homework.^49^ However, it is important to note that engagement in our study mostly refers expressing gratitude and asking for help vs completing homework and attending therapy sessions, which are related concepts.

We also found that most demographic and clinical baseline characteristics were unrelated with engagement with *CuidaTEXT*. This finding is in line with the literature on technological adjuncts to increase client adherence to psychotherapy, which suggests that baseline characteristics like symptom severity, educational background, or levels of coping skills are not associated with engagement between therapy sessions.^46^ The lack of a profile for frequent vs less frequent engagement caregivers suggests that there is no need to adapt the intervention to increase engagement and potentially intervention acceptability, among specific groups.

Compared to two studies assessing the effectiveness and user experiences of text messaging interventions, Latinos participating in this study exhibited a higher level of interaction, sending on average 83.95 text messages over a six-month period. Cartujana- Barrera et al (2019) found that among a mostly Latino sample of smokers, those who interacted with the program at least once sent on average 31.8 text messages during a 12-week period.

Sosa et al. (2017) found that among a mostly non-Hispanic sample of head and neck cancer patients, participants sent 12 text messages during a one-week period.^50^ Much like the study conducted with Latino smokers, participants in this study showed a preference for composing their own text messages rather than relying on predefined keywords from the intervention for a response. This implies that depending solely on keywords may not provide adequate support for Latino caregivers through text messaging. Therefore, there might be supplementary expenses associated with deploying trained personnel to respond to participants’ text messages, as observed in our study.

This study had several limitations that should be considered when interpreting the findings. This study included a small sample size (n=24) and did not have a control group, which hinders our ability to establish causal relationships between the intervention and observed outcomes. Follow-up was limited to a single assessment post-intervention at 6 months, which prevent from assessing directionality of associations. The composition of the sample included primarily middle-aged Latino women with some college education, who were already familiar with text messages. While the most common profile of Latino family caregivers of people with ADRD is the daughter, our sample could have included a larger number of men and spouses, which might differ in their engagement with caregiver support interventions and text messaging. Lastly, neither the participants, the assessment staff, nor the data analyst were blinded, potentially introducing bias into the results.

The study has implications for clinical practice and research. With regards to clinical practice, it is encouraging to see that while *CuidaTEXT* might lead to the reduction in depressive symptoms and other clinical outcomes,^18^ caregivers’ level of engagement is not associated to that success. The association of engagement with acceptability outcomes suggests that interventions could be planned to increase caregivers’ willingness to engage, to increase their satisfaction and related outcomes. Researchers can conduct qualitative work to understand how to best encourage engagement without adding burden to caregivers. The lack of association between engagement and most baseline variables, but existence of an association with acceptability outcomes suggests that people from diverse profiles may experience satisfaction and acceptability in general with the intervention. Also, the domains and examples of participants’ text messages found in our qualitative analyses can inform the development of future daily automatic or keyword messages, or a section for coaches on how to respond to specific messages in a protocolized way. Some interventions might propose more specialized answers to these messages, whereas others might refer caregivers to a resource who can help them responds to their needs. With regards to future research, text message studies on caregiving and other health and social issues should conduct similar research to explore whether their findings are comparable to ours. Research should include additional assessment points to explore the directionality of associations. For example, in our study we found an association between engagement with *CuidaTEXT* and acceptability outcomes (e.g., satisfaction with the intervention). However, it remains unclear whether engagement drove the satisfaction or those who were more satisfied engaged more in sending text messages to *CuidaTEXT.* Including mediators of these associations may also help understand this relationship. Finally, the lack of association between engagement and change in clinical outcomes reduces the chance for bias related to differential engagement with the intervention in a future randomized controlled study.

Text messaging provides a unique opportunity to increase access and improve clinical outcomes among Latino family caregivers of people with ADRD. However, little is known about how features of these interventions such as engagement with their text messages can benefit them. In this study we found that engagement with *CuidaTEXT*, was generally associated with acceptability outcomes, but not with baseline characteristics nor change in clinical outcomes. Encouraging engagement with caregiver support interventions might lead to higher levels of satisfaction with the intervention. However, studies need to explore directionality of these associations.

## Clinical Implications

- While *CuidaTEXT* shows promise in reducing depressive symptoms independently of caregiver engagement, the association between engagement and acceptability outcomes underscores the importance of planning interventions to enhance caregivers’ willingness to engage for increased satisfaction and related outcomes.
- Future researchers may consider the clinical significance of customizing interventions based on caregiver engagement and preferences, drawing on insights from qualitative analyses.

## Data Availability

None

---

**Appendix 1.**
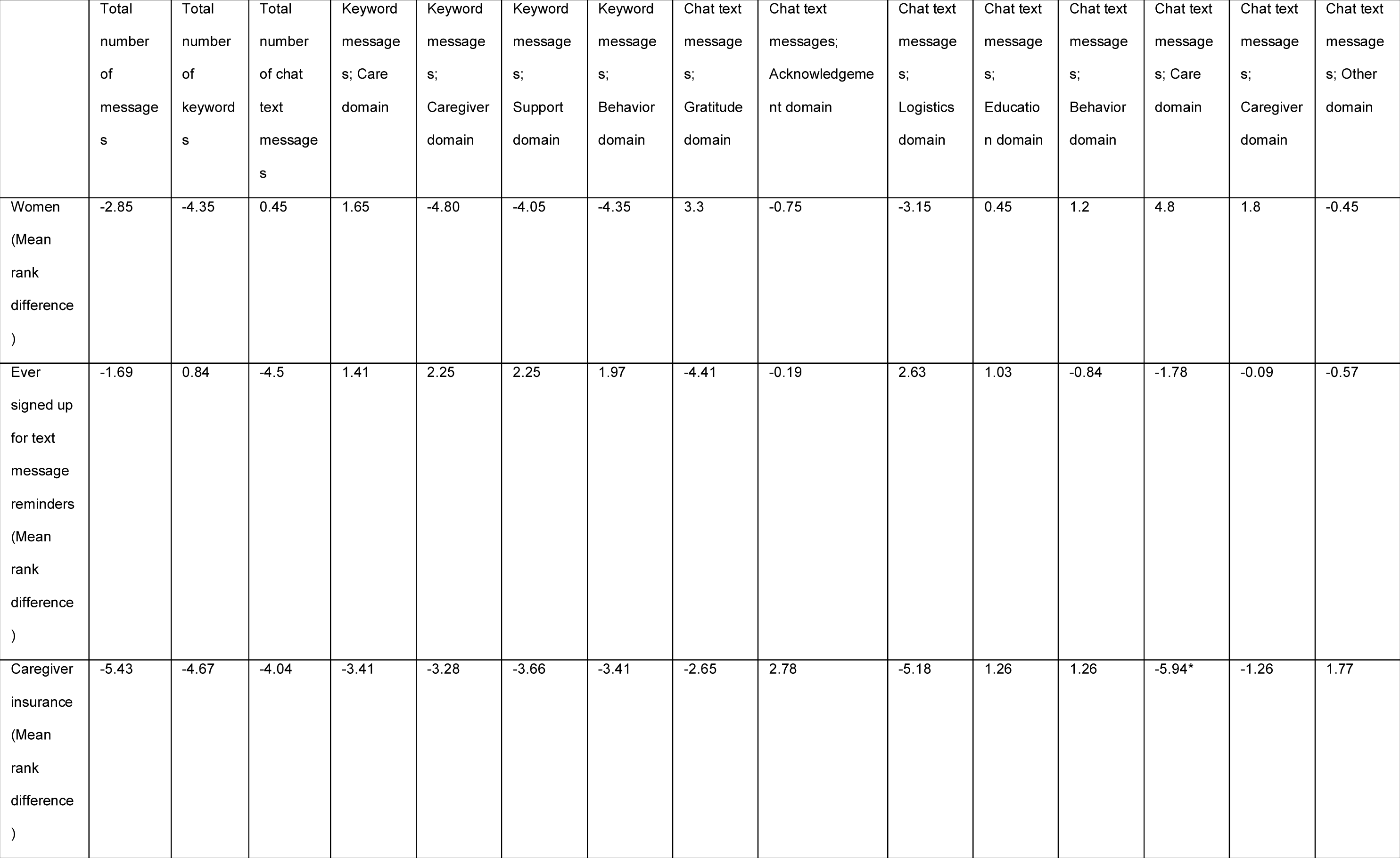
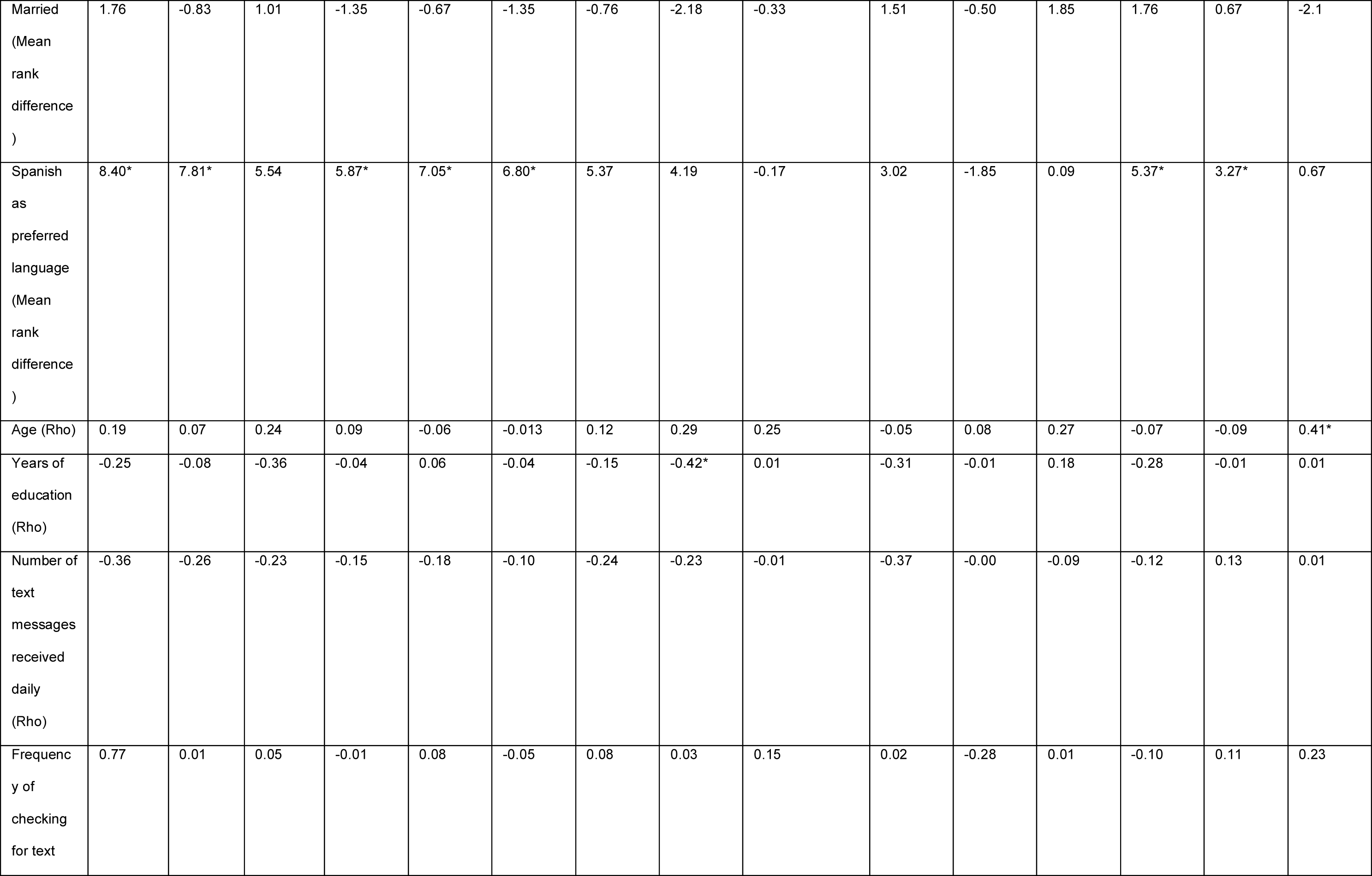
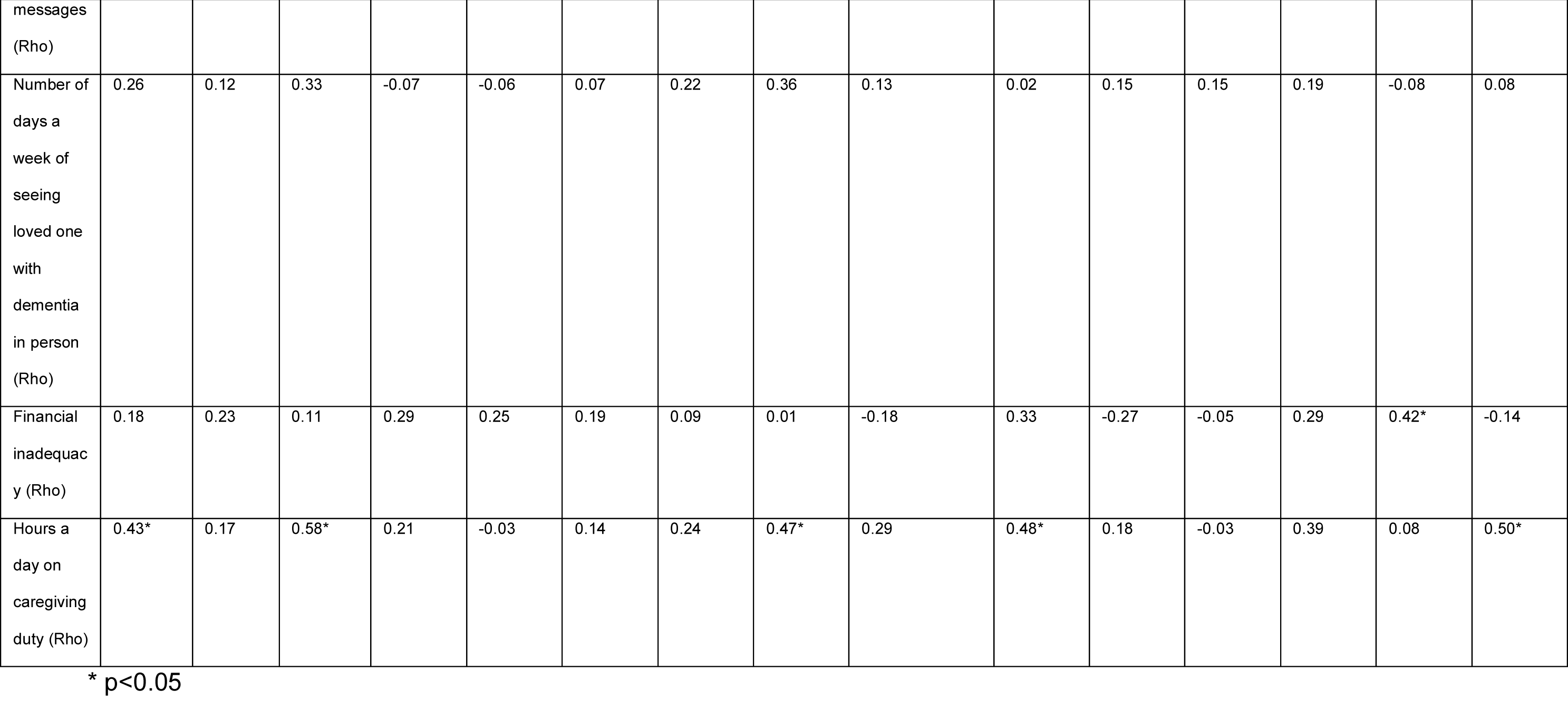
Association between engagement with CuidaTEXT and sociodemographic characteristics.

**Appendix 2.** Association between engagement with *CuidaTEXT* and baseline clinical characteristics.

|  | Total number of messages | Total number of keywords | Total number of chat text messages | Keyword messages; Care domain | Keyword messages; Caregiver domain | Keyword messages; Support domain | Keyword messages; Behavior domain | Chat text messages; Gratitude domain | Chat text messages; Acknowledgement domain | Chat text messages; Logistics domain | Chat text messages; Education domain | Chat text messages; Behavior domain | Chat text messages; Care domain | Chat text messages; Caregiver domain | Chat text messages; Other domain |
| --- | --- | --- | --- | --- | --- | --- | --- | --- | --- | --- | --- | --- | --- | --- | --- |
| NPI-Q Distress (Rho) | -0.076 | -0.070 | 0.021 | 0.074 | -0.076 | -0.067 | 0.097 | 0.14 | 0.06 | -0.06 | -0.17 | 0.03 | 0.22 | 0.33 | -0.32 |
| Caregiver Strain Index (Rho) | 0.224 | 0.085 | 0.365 | 0.299 | -0.046 | 0.080 | 0.006 | 0.31 | 0.12 | 0.15 | -0.22 | -0.05 | 0.43* | 0.39 | 0.08 |
| ZBI Burden (Rho) | -0.129 | -0.244 | 0.002 | -0.076 | -0.225 | -0.239 | -0.279 | -0.03 | -0.07 | 0.08 | -0.15 | -0.13 | 0.08 | 0.12 | -0.17 |
| Positive aspects of caregiving (Rho) | -.406* | -0.357 | -0.246 | -0.219 | -.467* | -0.280 | -0.280 | -0.13 | -0.07 | -0.00 | -0.04 | -0.01 | -0.26 | -0.28 | 0.33 |
| EEDS dementia knowledge (Rho) | -0.174 | -0.128 | -0.016 | 0.022 | -0.098 | -0.028 | -0.082 | -0.06 | 0.19 | 0.18 | 0.27 | 0.43* | -0.03 | 0.00 | 0.30 |
| ISEL-12 appraisal support (Rho) | -0.158 | -0.240 | -0.147 | -0.277 | -0.317 | -0.292 | -0.218 | -0.05 | -0.06 | -0.17 | 0.11 | -0.16 | -0.25 | -0.30 | -0.20 |
| ISEL-12<br>belonging<br>support<br>(Rho) | -0.05 | -0.38 | 0.04 | -.44* | -0.31 | -0.37 | -0.31 | 0.11 | 0.19 | -0.10 | 0.26 | -0.18 | -0.30 | -0.33 | -0.20 |
| ISEL-12<br>tangible<br>support<br>(Rho) | -0.14 | -0.06 | -0.12 | -0.16 | 0.00 | -0.10 | -0.15 | -0.08 | -0.06 | -0.24 | 0.16 | -0.15 | -0.22 | -0.17 | -0.33 |
| COPE 28<br>problem<br>focused<br>(Rho) | 0.27 | 0.16 | 0.16 | 0.14 | 0.21 | 0.31 | 0.09 | -0.07 | 0.22 | 0.03 | -0.15 | 0.23 | 0.22 | 0.38 | -0.09 |
| COPE 28<br>emotion<br>focused<br>(Rho) | 0.24 | 0.06 | 0.20 | 0.12 | 0.01 | 0.08 | 0.03 | 0.18 | 0.34 | -0.11 | -0.05 | -0.28 | 0.02 | 0.16 | 0.32 |
| COPE 28<br>avoidant<br>focused<br>(Rho) | -0.02 | -0.18 | 0.01 | -0.03 | -0.22 | -0.12 | -0.11 | -0.03 | -0.05 | 0.09 | -0.24 | -0.27 | 0.12 | 0.06 | -0.25 |
| CES-D-10<br>depression<br>(Rho) | -0.14 | -0.18 | -0.07 | -0.07 | -0.15 | -0.16 | -0.24 | -0.27 | -0.09 | 0.25 | -0.01 | -0.12 | 0.10 | -0.04 | -0.30 |
| SPANE-Positive<br>affect<br>(Rho) | 0.32 | 0.33 | 0.19 | 0.30 | 0.15 | 0.39 | 0.36 | 0.13 | -0.01 | 0.11 | -0.21 | 0.07 | 0.28 | 0.16 | 0.10 |
| SPANE-Negative<br>affect<br>(Rho) | -0.31 | -0.09 | -0.23 | 0.00 | -0.09 | 0.03 | -0.13 | -0.34 | -0.20 | 0.09 | -0.39 | -0.03 | 0.05 | 0.26 | -0.01 |
| PCS<br>Competenc<br>e (Rho) | -0.15 | -0.17 | -0.18 | -0.13 | -0.14 | -0.17 | -0.12 | -0.14 | 0.21 | -0.12 | -0.11 | 0.22 | -0.28 | -0.12 | -0.08 |
| Perceived<br>health<br>(Rho) | 0.20 | 0.36 | -0.11 | 0.27 | 0.35 | 0.35 | 0.40 | -0.13 | 0.10 | -0.24 | 0.07 | 0.26 | 0.06 | 0.12 | 0.21 |

\* $p < 0.05$

**Appendix 3.**
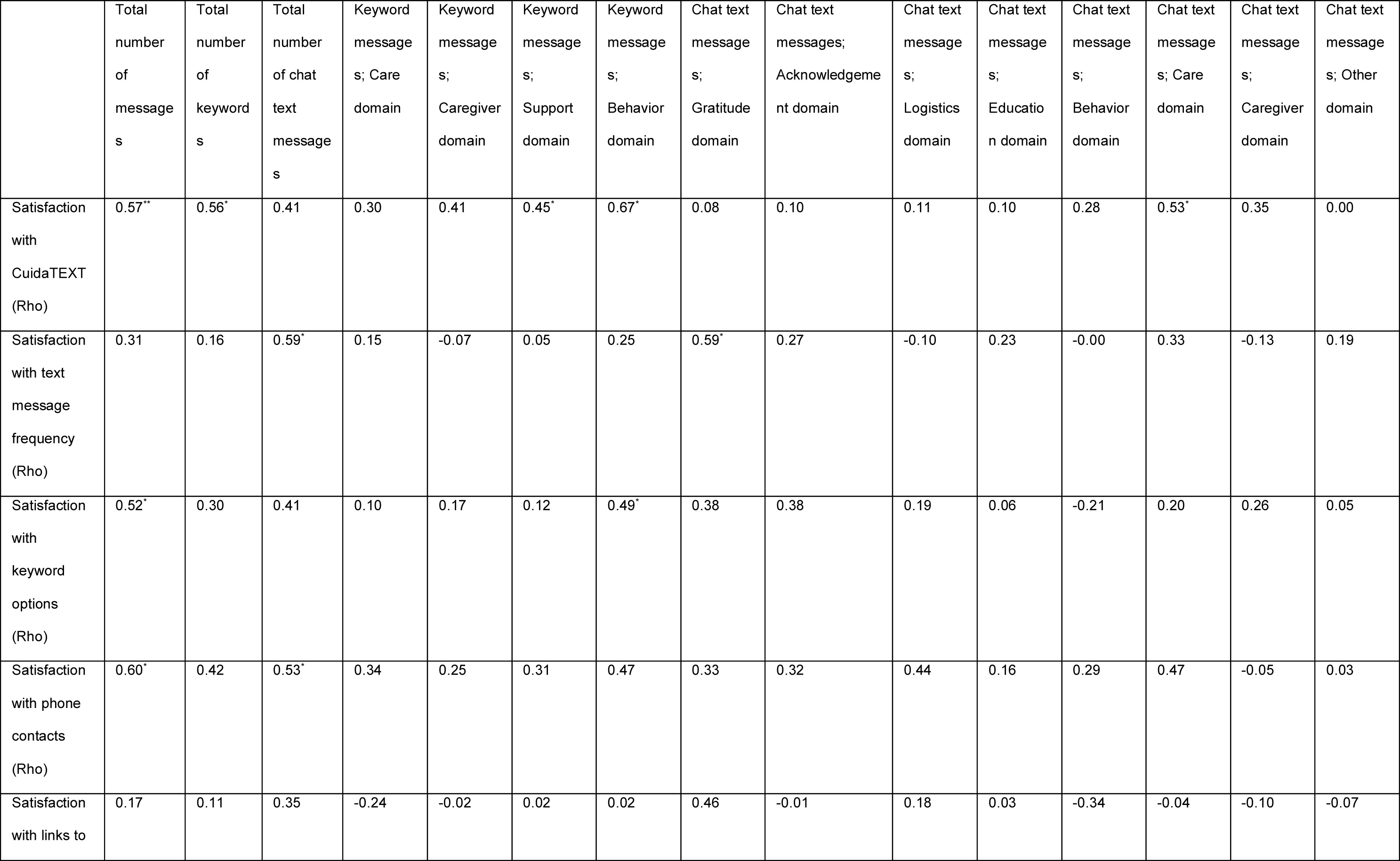
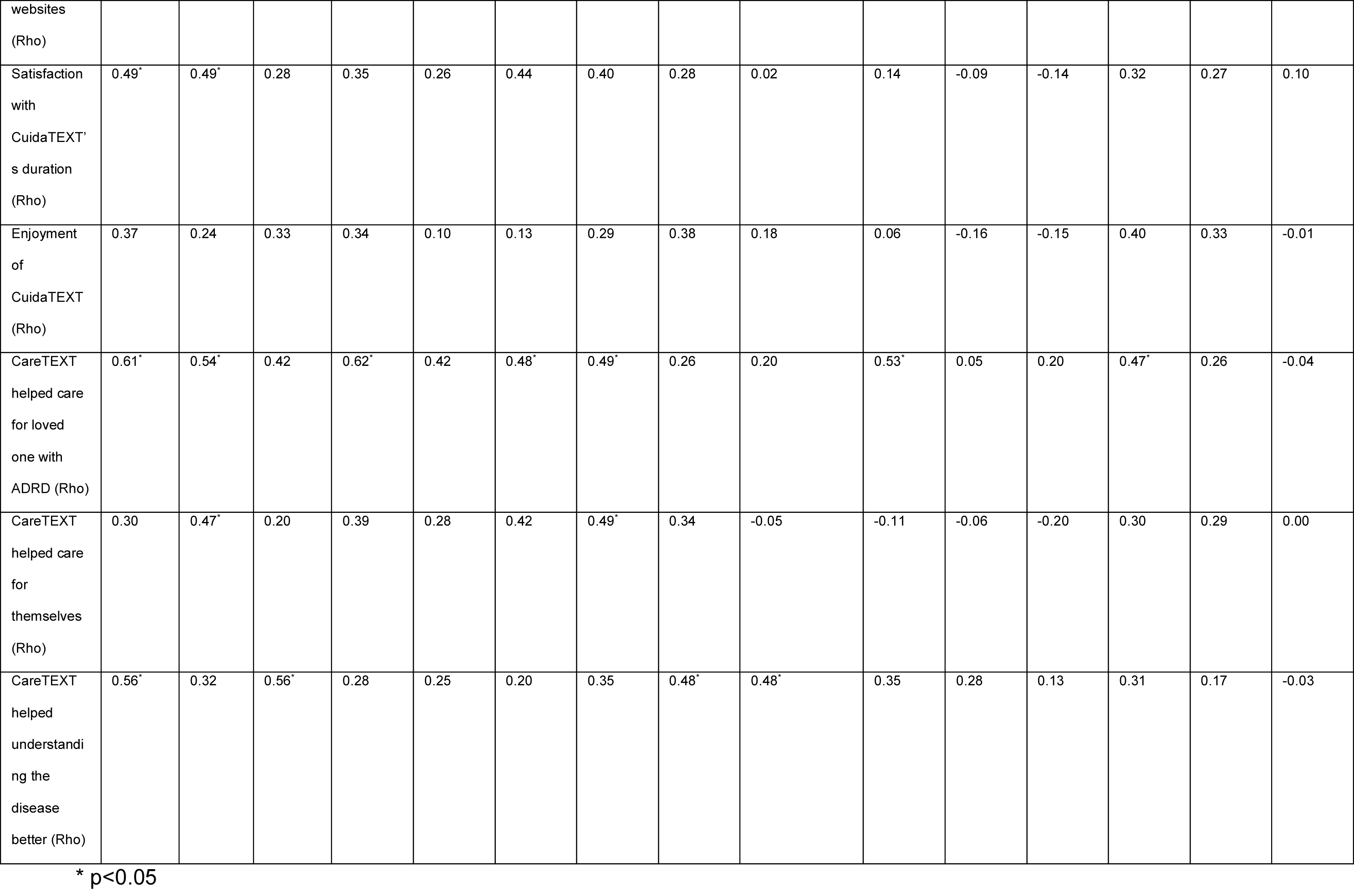
Association between engagement with *CuidaTEXT* and acceptability outcomes.

**Appendix 4.** Association between engagement with CuidaTEXT and baseline to follow-up change in clinical characteristics.

|  | Total number of messages | Total number of keywords | Total number of chat text messages | Keyword messages; Care domain | Keyword messages; Caregiver domain | Keyword messages; Support domain | Keyword messages; Behavior domain | Chat text messages; Gratitude domain | Chat text messages; Acknowledgement domain | Chat text messages; Logistics domain | Chat text messages; Education domain | Chat text messages; Behavior domain | Chat text messages; Care domain | Chat text messages; Caregiver domain | Chat text messages; Other domain |
| --- | --- | --- | --- | --- | --- | --- | --- | --- | --- | --- | --- | --- | --- | --- | --- |
| NPI-Q Distress (Rho) | 0.43 | 0.37 | 0.06 | 0.28 | 0.31 | 0.31 | 0.32 | -0.15 | 0.07 | -0.12 | -0.02 | 0.25 | 0.19 | 0.20 | 0.33 |
| Caregiver Strain Index (Rho) | 0.04 | -0.12 | 0.16 | -0.04 | -0.19 | 0.02 | -0.14 | -0.01 | 0.13 | 0.35 | -0.02 | 0.22 | - | - | - |
| ZBI Burden (Rho) | 0.07 | -0.03 | 0.13 | 0.15 | 0.01 | 0.10 | -0.07 | -0.02 | 0.11 | 0.45<sup>*</sup> | -0.10 | -0.00 | 0.11 | 0.02 | 0.28 |
| Positive aspects of caregiving (Rho) | 0.25 | 0.23 | 0.36 | 0.30 | 0.15 | 0.13 | 0.13 | 0.21 | -0.02 | 0.01 | 0.00 | 0.11 | 0.56<sup>*</sup> | 0.32 | -0.47<sup>*</sup> |
| EEDS dementia knowledge (Rho) | 0.18 | 0.17 | -0.04 | 0.05 | 0.14 | 0.10 | 0.12 | -0.13 | -0.18 | -0.05 | -0.12 | -0.21 | 0.20 | 0.12 | -0.50<sup>*</sup> |
| ISEL-12 appraisal support (Rho) | -0.23 | 0.10 | -0.17 | 0.14 | 0.06 | 0.19 | 0.03 | -0.15 | -0.23 | -0.13 | -0.17 | -0.23 | 0.08 | 0.28 | 0.19 |
| ISEL-12<br>belonging<br>support<br>(Rho) | -0.20 | -0.04 | -0.06 | 0.01 | -0.18 | -0.04 | 0.10 | 0.00 | -0.11 | -0.27 | -0.38 | 0.39 | 0.18 | 0.34 | 0.04 |
| ISEL-12<br>tangible<br>support<br>(Rho) | -0.15 | -0.10 | 0.02 | -0.08 | -0.25 | -0.07 | 0.06 | 0.12 | -0.13 | 0.03 | -0.18 | 0.23 | 0.17 | 0.18 | 0.01 |
| COPE 28<br>problem<br>focused<br>(Rho) | -0.16 | -0.13 | -0.15 | 0.03 | -0.16 | -0.25 | -0.06 | -0.12 | -0.19 | 0.31 | 0.08 | 0.09 | -0.03 | -0.45<sup>+</sup> | -0.15 |
| COPE 28<br>emotion<br>focused<br>(Rho) | -0.10 | -0.10 | -0.08 | 0.10 | -0.05 | -0.08 | -0.04 | -0.18 | -0.20 | 0.51<sup>+</sup> | 0.25 | 0.11 | 0.16 | -0.29 | -0.03 |
| COPE 28<br>avoidant<br>focused<br>(Rho) | 0.52<sup>+</sup> | 0.26 | 0.29 | 0.36 | 0.30 | 0.19 | 0.34 | 0.26 | 0.56<sup>+</sup> | 0.19 | 0.22 | 0.35 | 0.06 | 0.04 | 0.35 |
| CES-D-10<br>depression<br>(Rho) | -0.00 | -0.01 | -0.12 | 0.07 | -0.01 | 0.06 | 0.17 | 0.14 | 0.23 | -0.18 | -0.23 | 0.10 | -0.15 | 0.36 | 0.52<sup>+</sup> |
| SPANE-Positive<br>affect<br>(Rho) | -0.33 | -0.16 | -0.25 | -0.21 | -0.02 | -0.29 | -0.19 | -0.13 | 0.01 | -0.26 | 0.09 | -0.06 | -0.32 | -0.20 | -0.23 |
| SPANE-Negative<br>affect<br>(Rho) | 0.10 | -0.00 | 0.16 | 0.12 | 0.02 | -0.01 | 0.06 | 0.27 | 0.17 | 0.00 | 0.57<sup>*</sup> | 0.09 | 0.11 | -0.13 | 0.42 |
| PCS<br>Competenc<br>e (Rho) | 0.37 | 0.55<sup>*</sup> | -0.00 | 0.35 | 0.60<sup>*</sup> | 0.51<sup>*</sup> | 0.36 | -0.29 | -0.30 | 0.08 | 0.06 | 0.06 | 0.34 | 0.16 | -0.56<sup>*</sup> |
| Perceived<br>health<br>(Rho) | -0.04 | -0.03 | 0.04 | -0.19 | -0.04 | 0.01 | -0.04 | -0.10 | -0.09 | 0.32 | 0.13 | -0.28 | -0.07 | 0.00 | -0.22 |

\* p<0.05

## References

1. National Academies of Sciences & Medicine. Families caring for an aging America. 2016. 0309448093.

2. Alzheimer’s Association. 2016 Alzheimer’s disease facts and figures. Alzheimer’s & Dementia. 2016;12(4):459-509.

3. Bertrand RM, Fredman L, Saczynski J. Are all caregivers created equal? Stress in caregivers to adults with and without dementia. Journal of Aging and Health. 2006;18(4):534–551.

4. Friedman EM, Shih RA, Langa KM, Hurd MD. US prevalence and predictors of informal caregiving for dementia. Health Affairs. 2015;34(10):1637–1641.

5. Kasper JD, Freedman VA, Spillman BC, Wolff JL. The disproportionate impact of dementia on family and unpaid caregiving to older adults. Health Affairs. 2015;34(10):1642–1649.

6. Moon H, Townsend AL, Dilworth-Anderson P, Whitlatch CJ. Predictors of discrepancy between care recipients with mild-to-moderate dementia and their caregivers on perceptions of the care recipients’ quality of life. American Journal of Alzheimer’s Disease & Other Dementias®. 2016;31(6):508–515.

7. Ory MG, Hoffman III RR, Yee JL, Tennstedt S, Schulz R. Prevalence and impact of caregiving: A detailed comparison between dementia and nondementia caregivers. The Gerontologist. 1999;39(2):177–186.

8. Pinquart M, Sörensen S. Ethnic differences in stressors, resources, and psychological outcomes of family caregiving: A meta-analysis. The Gerontologist. 2005;45(1):90–106.

9. Feldman SJ, Solway E, Kirch M, Malani P, Singer D, Roberts JS. Correlates of formal support service use among dementia caregivers. Journal of gerontological social work. 2021;64(2):135–150.

10. Chen C, Thunell J, Zissimopoulos J. Changes in physical and mental health of Black, Hispanic, and White caregivers and non-caregivers associated with onset of spousal dementia. Alzheimer’s & Dementia: Translational Research & Clinical Interventions. 2020;6(1):e12082.

11. Scharlach AE, Giunta N, Chow JC-C, Lehning A. Racial and ethnic variations in caregiver service use. Journal of Aging and Health. 2008;20(3):326–346. doi:10.1177/0898264308315426

12. Pew Research Center. Mobile fact sheet. 2021.

13. Duggan M. *Cell phone activities* 2013. 2013. September 2013.

14. Schilling L, Bennett G, Bull S, Kempe A, Wretling M, Staton E. Text messaging in healthcare research toolkit. *Center for Research in Implementation Science and Prevention (CRISP)*, University of Colorado School of Medicine. 2013;

15. Hall AK, Cole-Lewis H, Bernhardt JM. Mobile text messaging for health: a systematic review of reviews. Annual review of public health. 2015;36:393–415.

16. Guerriero C, Cairns J, Roberts I, Rodgers A, Whittaker R, Free C. The cost-effectiveness of smoking cessation support delivered by mobile phone text messaging: Txt2stop. Eur J Health Econ. Oct 2013;14(5):789–97. doi:10.1007/s10198-012-0424-5

17. Zurovac D, Larson BA, Sudoi RK, Snow RW. Costs and cost-effectiveness of a mobile phone text-message reminder programmes to improve health workers’ adherence to malaria guidelines in Kenya. PLoS One. 2012;7(12):e52045.

18. Perales-Puchalt J, Ramírez-Mantilla M, Fracachán-Cabrera M, et al. A text message intervention to support latino dementia family caregivers (CuidaTEXT): feasibility study. Clinical Gerontologist. 2022:1–16. doi:10.1080/07317115.2022.2137449

19. Perales-Puchalt J, Acosta-Rullán M, Ramírez-Mantilla M, et al. A Text Messaging Intervention to Support Latinx Family Caregivers of Individuals With Dementia (CuidaTEXT): Development and Usability Study. JMIR aging. 2022;5(2):e35625.

20. Pearlin LI, Mullan JT, Semple SJ, Skaff MM. Caregiving and the stress process: An overview of concepts and their measures. The Gerontologist. 1990;30(5):583–594.

21. Bandura A. Social foundations of thought and action: a social cognitive theory. Englewood Cliffs, NJ. 1986;1986

22. Cartujano-Barrera F, Sanderson Cox L, Arana-Chicas E, et al. Feasibility and acceptability of a culturally-and linguistically-adapted smoking cessation text messaging intervention for Latino smokers. Frontiers in Public Health. 2020;8:269.

23. Abroms L, Hershcovitz R, Boal A, Levine H. Feasibility and Acceptability of a Text Messaging Program for Smoking Cessation in Israel. J Health Commun. Aug 2015;20(8):903–9. doi:10.1080/10810730.2015.1018585

24. Acevedo A, Krueger KR, Navarro E, et al. The Spanish translation and adaptation of the uniform data set of the National Institute on Aging Alzheimer’s Disease Centers. Alzheimer disease and associated disorders. 2009;23(2):102.

25. Kaufer DI, Cummings JL, Ketchel P, et al. Validation of the NPI-Q, a brief clinical form of the Neuropsychiatric Inventory. The Journal of neuropsychiatry and clinical neurosciences. 2000;12(2):233–239.

26. Thornton M, Travis SS. Analysis of the reliability of the modified caregiver strain index. The Journals of Gerontology Series B: Psychological Sciences and Social Sciences. 2003;58(2):S127–S132.

27. Higginson IJ, Gao W, Jackson D, Murray J, Harding R. Short-form Zarit Caregiver Burden Interviews were valid in advanced conditions. Journal of clinical epidemiology. 2010;63(5):535–542.

28. Tarlow BJ, Wisniewski SR, Belle SH, Rubert M, Ory MG, Gallagher-Thompson D. Positive aspects of caregiving: Contributions of the REACH project to the development of new measures for Alzheimer’s caregiving. Research on aging. 2004;26(4):429–453.

29. Connell C, Holmes S. Knowledge, attitudes, and beliefs about Alzheimer’s disease. 1996:

30. Roberts JS, Connell CM. Illness representations among first-degree relatives of people with Alzheimer disease. Alzheimer Disease & Associated Disorders. 2000;14(3):129–136.

31. Merz EL, Roesch SC, Malcarne VL, et al. Validation of interpersonal support evaluation list-12 (ISEL-12) scores among English-and Spanish-speaking Hispanics/Latinos from the HCHS/SOL Sociocultural Ancillary Study. Psychological assessment. 2014;26(2):384.

32. Carver CS. You want to measure coping but your protocol’too long: Consider the brief cope. International journal of behavioral medicine. 1997;4(1):92–100.

33. Perczek R, Carver CS, Price AA, Pozo-Kaderman C. Coping, mood, and aspects of personality in Spanish translation and evidence of convergence with English versions. Journal of personality Assessment. 2000;74(1):63–87.

34. Cheng ST, Chan AC. The center for epidemiologic studies depression scale in older Chinese: thresholds for long and short forms. International Journal of Geriatric Psychiatry: A journal of the psychiatry of late life and allied sciences. 2005;20(5):465–470.

35. González P, Nuñez A, Merz E, et al. Measurement properties of the Center for Epidemiologic Studies Depression Scale (CES-D 10): Findings from HCHS/SOL. Psychological assessment. 2017;29(4):372.

36. Diener E, Wirtz D, Tov W, et al. New well-being measures: Short scales to assess flourishing and positive and negative feelings. Social Indicators Research. 2010;97(2):143–156.

37. Daniel-González L, de la Rubia JM, Valle de la O A, Garcia-Cadena CH, Martinez-Marti ML. Validation of the mexican spanish version of the scale of positive and negative experience in a sample of medical and psychology students. Psychological Reports. 2020;123(5):2053–2079.

38. Carter JH, Stewart BJ, Archbold PG, et al. Living with a person who has Parkinson’s disease: the spouse’s perspective by stage of disease. Movement disorders: official journal of the Movement Disorder Society. 1998;13(1):20–28.

39. Gutierrez-Baena B, Romero-Grimaldi C. Development and psychometric testing of the Spanish version of the Caregiver Preparedness Scale. Nursing Open. 2021;8(3):1183–1193.

40. Patel KV, Eschbach K, Rudkin LL, Peek MK, Markides KS. Neighborhood context and self-rated health in older Mexican Americans. Annals of epidemiology. 2003;13(9):620–628.

41. Basch CE. Focus group interview: an underutilized research technique for improving theory and practice in health education. Health education quarterly. 1987;14(4):411–448. doi:10.1177/109019818701400404

42. Neergaard MA, Olesen F, Andersen RS, Sondergaard J. Qualitative description–the poor cousin of health research? BMC medical research methodology. 2009;9(1):1–5. doi:10.1186/1471-2288-9-52

43. Miles MB, Huberman AM. Qualitative data analysis: A sourcebook of new methods. Qualitative data analysis: a sourcebook of new methods. 1984:263–263.

44. IBM Corp. Released 2013. IBM SPSS Statistics for Windows, Version 22.0.: Armonk, NY: IBM Corp.; 2013.

45. Hebert C. *Examining text messaging frequency, interpersonal communication skills, and relational satisfaction among college-age students: A correlational study*. Northcentral University; 2016.

46. Clough BA, Casey LM. Technological adjuncts to increase adherence to therapy: a review. Clinical psychology review. 2011;31(5):697–710.

47. Loeb KL, Wilson GT, Labouvie E, et al. Therapeutic alliance and treatment adherence in two interventions for bulimia nervosa: a study of process and outcome. Journal of consulting and clinical psychology. 2005;73(6):1097.

48. Abramowitz JS, Franklin ME, Zoellner LA, Dibernardo CL. Treatment compliance and outcome in obsessive-compulsive disorder. Behavior modification. 2002;26(4):447–463.

49. Burns DD, Spangler DL. Does psychotherapy homework lead to improvements in depression in cognitive–behavioral therapy or does improvement lead to increased homework compliance? Journal of consulting and clinical psychology. 2000;68(1):46.

50. Sosa A, Heineman N, Thomas K, et al. Improving patient health engagement with mobile texting: a pilot study in the head and neck postoperative setting. Head & neck. 2017;39(5):988–995.

